# A Utility-Based Machine Learning-Driven Personalized Lifestyle Recommendation for Cardiovascular Disease Prevention

**DOI:** 10.1101/2022.02.02.22270339

**Authors:** Ayse Dogan, Yuxuan Li, Chiwetalu Peter Odo, Kalyani Sonawane, Ying Lin, Chenang Liu

## Abstract

In recent decades, cardiovascular disease (CVD) has become the leading cause of death in most countries of the world. Since many types of CVD are preventable by modifying lifestyle behaviors, the objective of this paper is to develop an effective personalized lifestyle recommendation algorithm for reducing the risk of common types of CVD. However, in practice, the underlying relationships between the risk factors (e.g., lifestyles, blood pressure, etc.) and disease onset is highly complex. It is also challenging to identify the effective modification recommendations for different individuals due to the uncertainties in individuals’ preference to change and disease progression. Therefore, to address these challenges, this study developed a novel data-driven approach for personalized lifestyle behaviors recommendation based on machine learning and a personalized exponential utility function model. The contributions of this work can be summarized into three aspects: (1) a classification-based prediction model is implemented to accurately predict the CVD risk based on the condition of risk factors; (2) the generative adversarial network (GAN) is incorporated to learn the underlying relationship between risk factors, as well as quantifying the uncertainty of disease progression under lifestyle modifications; and (3) a novel personalized exponential utility function model is proposed to evaluate the modifications’ utilities with respect to CVD risk reduction, individual’s preference to change and disease progression uncertainty, and identify the optimal modification for each individual. The effectiveness of the proposed method is validated through an open-access CVD dataset. The results demonstrate that the personalized lifestyle modification recommended by the proposed methodology can significantly reduce the potential CVD risk. Thus, it is very promising to be further applied to real-world cases for CVD prevention.

## 1 Introduction

The objective of this study is to develop an effective data-driven personalized lifestyle recommendation algorithm for reducing the individual’s cardiovascular disease (CVD) risk. CVD is a class of common chronic diseases that affect the heart or blood vessel functions, such as myocardial infarction (MI), stroke, and congenital heart defect (CHD). It has been the most frequent cause of death in the United States for a long time [1], e.g., sudden cardiac death. Although CVD poses a great challenge for human health, existing studies have shown that most types of CVD could be potentially well-prevented if the related risk factors can be controlled appropriately, particularly, for the lifestyle behaviors, such as tobacco, diet, and exercise [2, 3].

Several evidence-based clincal guidelines, have been developed for the CVD prevention, such as the lifestyle modification recommendations in AHA Scientific Statement [4]. In general, they are population-orientied lifestyle recommendation protocols, which aim to reduce the CVD risk over the whole population [4]. For example, the clinical guidelines recommend specific thresholds of diet and lifestyle, such as limit dietary cholesterol to be lower than 300 mg per day, and control total carbohydrate intake between 45% and 65% of energy [5]. However, these generalized recommendations fail to consider individuals’ particular characteristics and preferences, and hence may perform poorly at personalized risk reduction and ensuring lifespan adherence to intervention plans [6]. Thus, to better prevent the CVD on vulnerable individuals, an effective methodology that is able to provide personalized suggestions for lifestyle behavior modification is critically needed.

Recently the enrichment of healthcare database enables the development of personalized lifestyle modification from a data-driven perspective. For example, a recent study utilized the Atherosclerosis Risk in Communities (ARIC) study [7], which integrates the survey data, clinical test, medical diagnosis, and population surveillance data, to develop a personalized recommendation approach using machine learning, and its results have also demonstrated the promising potentials of this direction [6]. However, to achieve satisfactory performance, there are still two major challenges in the development of personalized lifestyle modification recommendation. First, the effects of lifestyle modification on CVD disease progression of each individual is difficult to predict due to the complex interactions between lifestyle factors (directly changeable), physiological symptoms (indirectly changeable), individual’s characteristics (unchangeable), and disease risk. As a minor change in lifestyle factor may lead to the progressions in multiple physiological symptoms and further affect the disease risk, it is critical to capture the interactions between risk factors and quantify the potential changes in other factors under the potential lifestyle modifications. Second, the individuals’ preferences to change the lifestyle behaviors were usually ignored in existing recommendation algorithms, which results in the low adherence and utility of recommended lifestyle modifications in reality. Therefore, an effective evaluation model for the personalized lifestyle modifications considering the individual’s preference to change as well as the modifications’ effectiveness and uncertainty on CVD risk reduction is needed.

To address these challenges, this paper proposed a new methodology by incorporating the emerging generative adversarial network (GAN) [8] and a novel personalized utility function model, with the consideration of individual’s preference to change and disease progression uncertainty, particularly, the uncertainties in the indirectly changable risk factors (i.e., physiological symptoms). The specific contribution of this work can be summarized by three aspects: (1) a classification-based prediction model is implemented to effectively predict the CVD risk based on the given condition of risk factors; (2) the powerful generative adversarial network (GAN) is successfully incorporated to learn the underlying relationship between risk factors, as well as quantifying the uncertainty of disease progression under lifestyle modifications; and (3) a novel personalized exponential utility function model is proposed to assess lifestyle modification in terms of the individual’s preference as well as the modification effectiveness and uncertainty, enabling to identify the optimal modification that achieves desired CVD risk reduction and individual preference.

To predict the individual’s CVD risk based on the given condition of risk factors, supervised machine learning technques are applied to quantify the risk factors and disease outcomes. Compared with the existing studies that only consider a pre-determined approach, this work further explored different popular algorithms, and then select the most accurate one through comparison.

Developing a GAN-based approach to model the interactions between risk factors is another key contribution of this work. These interactions are highly complex and contains significant uncertainties, particularly, for the physiological symptoms related factors, which are indirectly changeable. However, existing approaches did not sufficiently considered these uncertainties in the modeling and analysis [6, 9]. Recently GAN has become one of the most popular machine learning models for learning the complex distribution of multivariate data. In the proposed method, GAN is able to generate applicable alternative lifestyle behaviors by considering the joint distribution of risk factors. Meanwhile, with the consideration of uncertainties, it is also applied to synthesize the progressions of the related physiological symptoms under the potential lifestyle modifications, and thereby enable effective CVD risk prediction for the alternative lifestyle behaviors.

More importantly, inspired by the expected utility theory, the developed utility function-based decision-making model also significantly improves the rational to identify the optimal modification, since it can jointly consider the individual’s CVD risk, preference to change, and disease progression uncertainty in the modification assessment, which have not been well addressed in the existing studies [6, 9].

The rest of this paper is organized as follows. A brief review of the research background and related work is provided in Sec. 2. The proposed research methodology is presented in detail in Sec. 3. Subsequently, the results are demonstrated in Sec. 4. Finally, conclusions and discussions are presented in Sec. 5.

## 2 Review of Related Work

This paper focuses on developing a data-driven lifestyle recommendation algorithm for reducing the risk of CVD. Thus, this section first briefly reviewed the related studies on the effect of lifestyle behavior on CVD (Sec. 2.1), then it is followed by a review of the existing data-driven recommendation approaches for disease prevention (Sec. 2.2).

### 2.1 Effect of lifestyle behaviors on the risk of CVDs

To prevent the onset of heart failure, the effects of lifestyle behaviors on CVD risk have become one of the most critical areas in heart disease research. Several popular long-term cohort studies regarding heart disease, including the Framingham heart study (FHS) [10] and Atherosclerosis Risk in Communities (ARIC) Study, provided great data resources and research opportunities to investigate the CVD risk factors. For example, through FHS, Vasan *et al*. [11] explored the correlation between antecedent blood pressure and the risk of CVD, and Wilson *et al*. [12] investigated the effect of blood pressure and cholesterol on CHD risk together. Regarding the long-term risk prediction, Pencina *et al*. [13] developed a statistical model using the FHS data to predict the CVD risks in 10 years and 30 years. Wickramasinghe *et al*. [14] further considered the fitness level for 30-year risk cardiovascular mortality prediction. Lloyd-Jones [15] also provided a review of the history and principles of CVD risk prediction. Furthermore, considering the effects of lifestyle behaviors, E Millen *et al*. [16] examined the relationship between dietary patterns, food frequency, and atherosclerotic disease by using the FHS. Recently, more studies were conducted based on the ARIC data, for example, Chi *et al*. [6], Mansoor *et al*. [17], and Chambless *et al*. [18] investigated the effect of different lifestyles on CVD risks. In these studies, different machine learning and statistical modeling methods were applied, including *k* nearest neighbors, Kaplan-Meier analysis, and Cox regression model, etc. Although the above-mentioned studies made great progress in the analysis of CVD risk in recent decades, it still lacks effective methods to provide individualized guidelines, particularly, lifestyle modification plans, for CVD prevention.

### 2.2 Data-driven recommendation approaches

Besides the risk prediction and assessment, it is also critical to provide personalized recommendations for health improvement, which has been investigated in several existing studies, including Enwald *et al*. [19], Skinner *et al*. [20], Kreuter *et al*. [21], Brug *et al*. [22], and Oenema *et al*. [23]. For example, Enwald *et al*. [19] presented that nutrition and physical exercises are supportive tailoring interventions, which have a positive health effect since the individuals could have a more active role in their health. However, these studies assumed that all the features can be directly affected by human intervention. Besides, the correlations between different features were also not considered.

Recently, data-driven recommendation approaches have become more and more popular in a large variety of areas, and they also demonstrate great potential for recommending alternative lifestyles to reduce the risk of diagnosing with diseases for individuals. For example, using the data provided by cardiologist, Jabeen *et al*. [24] developed an approach for CVD treatment suggestion based on a perspective of the Internet of Things. Another perspective, using the ARIC database, Chi *et al*. [6] developed an expert system for lifestyle recommendations to reduce the CVD risk using the *k*-NN algorithm. Nam *et al*. [25] also developed an expert system for monitoring physical activity and recommending lifestyle interventions in obesity. The developed system is able to output a diet and exercise program for treating obesity. However, the above-mentioned studies did not sufficiently consider the effects of disease progressions and individual’s preference to change lifestyles, which usually play critical roles in practice.

Among various data-driven recommendation methodologies, as an emerging technique, the generative adversarial network (GAN) recently becomes one of the most famous choices. As a powerful generative model in machine learning, GAN aims to synthesize potential recommendations through adversarial learning [26]. Its strong capability to learn the distribution of complex real data offers great potentials in its applications to recommendation systems. Particularly, it is promising to address two common limitations that many existing methods suffer from, namely, the data noise issue and data sparsity issue [27]. Thus, the GAN-based recommendation algorithms have been developed and applied in many fields with different recommendation types, such as Kang *et al*. [28], He *et al*. [29], and Li *et al*. [30]. Furthermore, recent studies also try to integrate GAN with other powerful machine learning techniques to further improve the recommendation accuracy and effectiveness. For example, the RecGAN [31] integrates the recurrent neural network (RNN) and GAN for the development of recommendation systems. The popular reinforcement learning (RL) model, which can imitate the user behaviors dynamically, is also effectively combined with GAN in the recommendation system [32]. Moreover, the DRCGR [33] further incoprates both neural networks and reinforcement learning (RL) with GAN in the interactive recommendation system. Although GAN has been successfully applied for these various recommendation systems, as a very powerful technique, its applications in lifestyle recommendation for reducing the disease risk, particularly, CVD risk, is still very limited.

## 3 Proposed Research Methodology

As shown in Figure 1, the overall framework of the proposed research methodology consists of three steps: (1) a cohort from the ARIC database is studied, and the features and outcome variables are defined (Sec. 3.1); (2) a classification-based predictive model is developed for individual CVD risk assessment (Sec. 3.2); and (3) a utility-based lifestyle recommendation algorithm is proposed to reduce the individual’s CVD risk by incorporating GAN and utility function model (Sec. 3.3). Additionally, the approaches to validate the effectiveness of the proposed methodology are described in Sec. 3.4.

**Figure 1:**
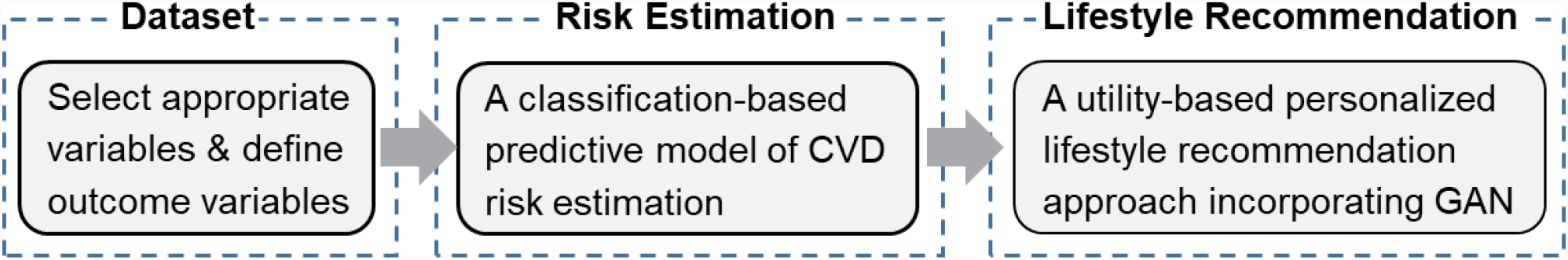
The overall framework of the proposed research methodology.

### 3.1 Data description

The data used in this study is a part of the Cohort Component of the Atherosclerosis Risk in Communities (ARIC) database [7], which was collected from four US communities: (1) Forsyth Country, NC; (2) Jackson, MS; (3) Suburban Minneapolis, MN; and (4) Washington Country, MD [18]. In each community, approximately 4,000 individuals aged 45-64 were randomly selected and recruited to collect their medical, social, and demographic data. In this study, the data collected from the participants’ first visit during 1987-1989 (i.e., visit 1) are used for the methodology development and validation. Meanwhile, participants who were already diagnosed with any type of heart failure or CVD before the first visit are removed. Therefore, a total of 13,654 participants are obtained. Subsequently, based on the literature [6], the risk factors that may impact the CVD risk are identified (listed in APPENDIX 1**Error! Reference source not found**.). Notably, in addition to the factors identified in [6], this study further includes the medication variables that are closed related to the occurance of CVDs. The selected variables are distinguished into three categories: directly changeable variables, indirectly changeable variables, and unchangeable variables.

The directly changeable variables describe each participant’s lifestyle behaviors, including diet, exercise, cigarette, alcohol, etc. In general, these variables can be directly changed by individuals’ lifestyle behaviors. As mentioned in Sec. 1, this study aims to identify the optimal modifications of each individual’s lifestyle behaviors, which is essential to appropriately adjust the directly changeable variables. In this study, 11 directly changeable variables (see more details in APPENDIX 1 Table 2) are selected from the ARIC database based on the existing literature [5, 6], which are denoted as,

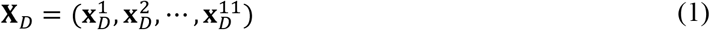

where 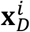 represents the *i*^th^ directly changeable variable. Considering the modification applicability in practice, the continuous variables in **X**_*D*_ are discretized to five intervals based on quantiles, while the categorical variables remain the same.

The indirectly changeable variables (14 variables in this study), denoted as **X**_*I*_, represent the physiological symptoms related to the health of participants, such as systolic and diastolic blood pressures, apolipoprotein, creatinine, etc. Notably, these variables can be affected by the changes of lifestyle behaviors. For instance, there is a strong causal relationship between blood pressure and the dietary such as the amount of salt consumption [34]. Therefore, participants can indirectly reduce or increase these variables by changing the directly changeable variables. Besides, there are several variables representing the characteristics of the individuals that cannot be changed, e.g., participant’s demographic information, height and medication history. Thus, these are named unchangeable variables, **X**_*U*_. In this study, there are in total 15 selected unchangeable variables.

Furthermore, four 10-year CVD outcomes (*y*_*i*_, *i* = 1,2,3,4) are defined using the Community Surveillance Component in ARIC, which indicate the occurrence of Fatal CHD (*y*_1_), MI (*y*_2_), and Stroke (*y*_3_), as well as any type of them (*y*_4_) within the 10 years follow-up.

### 3.2 Classification model for CVD risk estimation

To identify the high-risk patients and evaluate the effectiveness of the suggested lifestyle modifications, the CVD risk of each participant needs to be accurately predicted using the selected risk-related variables (**X**_*I*_, **X**_*U*_, **X**_*D*_), i.e., to develop a predictive model, as formulated in Eq. (2),

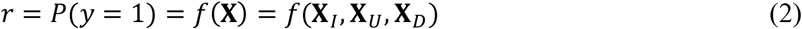

where *f* is an unknown nonlinear function to predict the CVD risk. To accurately predict CVD outcomes, a set of classification algorithms are applied to estimate the predictive model for each outcome individually.

In practice, due to several practical reasons, for example, rejected responses to some survey questions or some unincluded test results, missing values exist in the collected dataset. Consequently, to ensure model accuracy, it is necessary to impute the missing values before training *f*. In this study, a widely applied data-driven imputation approach, namely, *k* nearest neighbor (*k*-NN) imputation, is applied after comparison. The idea of *k*-NN imputation is to estimate the missing value by *k* assigned samples (nearest neighbors) that are similar or close in the dataset and then it can be imputed using the mean value of these *k* neighbors in the dataset [35]. In practice, the value of *k* plays a significant role since a smaller *k* may cause underfitting while a larger one may result in overfitting. Therefore, different *k* values are tried and the one that provides the best performance is selected in the study.

Subsequently, the widely-applied classification algorithms, including random forest (RF) [36], support vector machine (SVM) [37], and *k*-NN classification [38], are thereby applied to fit *f* after missing value imputation. For model comparison and selection, the receiver operating characteristic (ROC) curve [39] and its AUC (area under the curve) score [39], are utilized. Through comparison (see more details in Section **Error! Reference source not found**.), the random forest is selected to fit a CVD risk prediction model 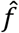 to approximate *f*. It is worth mentioning that the resampling technique is also incorporated when training the classfiers, which helps to reduce the effects from the data imbalanced issue.

### 3.3 Personalized lifestyle recommendation using GAN and regularized utility function

Based on the CVD risk prediction model developed in Sec. 3.2, each patient’s expected risk after a lifestyle modification can be predicted. However, there are still two major challenges to identify the optimal lifestyle modification for each patient: (1) the complex interactions between **X**_*I*_,, **X**_*U*_, and **X**_*D*_ pose great uncertainty in disease progression; and (2) the willingness and cost to change lifestyles of the patients should be considered as well. To address these two challenges, as illustrated in Figure 2, a novel personalized lifestyle recommendation algorithm is proposed by integrating the GAN method and a new regularized expected utility function model. Specifically, patients with high predicted risk of CVD are considered for lifestyle modification recommendation. For each high risk patient, a set of applicable lifestyle modifications and the associated changes in indirectly changeable factors are generated from the GAN method conditioning on patient’s unchangeable factor profiles. Then the expected utility of each applicable lifestyle modification is further evaluated through a regularized utility function model in the context of patients’ willingness and cost to change lifestyles. An optimal lifestyle modification with the highest expected utility value is finally selected for the patient’s personalized lifestyle modification recommendation.

**Figure 2:**
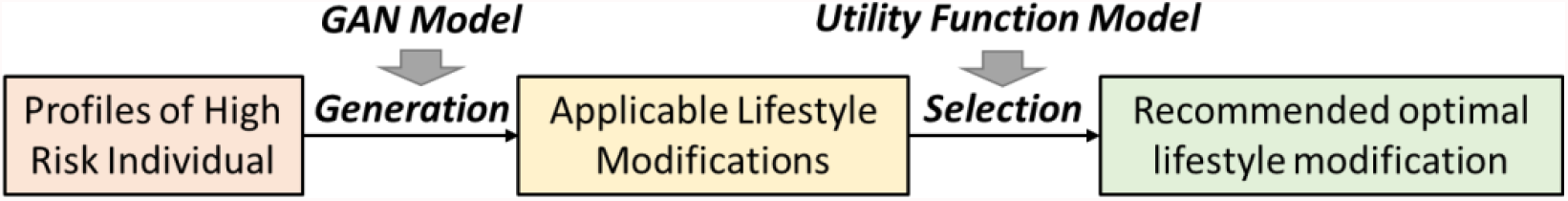
The overall framework of the proposed personalized lifestyle recommendation approach.

#### 3.3.1 Generative adversarial networks (GAN)

As a popular emerging machine learning model, GAN has demonstrated its strong capability of capturing the complex underlying relationship between variables. The main idea of GAN [8] is to train two networks, namely, generator *G* and discriminator *D*, with a minimax game for *D* and *G* demonstrated in Eq. (3),

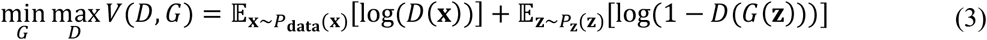

where the generator *G* generates artificial samples **z** and the discriminator *D* aims to label the input samples **x**. *G* is trying to trick the *D* by its artificial samples and make *D* be unable to distinguish whether it is artificial or actual. In another word, *G* is aiming to produce artificial data that is similar to original ones.

In this study, an extension of GAN, namely, the generative adversarial imputation networks (GAIN) [40], is applied to implement the following two tasks: (1) generate applicable alternative lifestyle behaviors (lifestyle modifications) for the high risk patients, i.e., generate alternative **X**_*D*_ by a given **X**_*U*_; and (2) generate feasible indirectly changeable variables associated with the generated alternative lifestyle behaviors for the high risk patients, i.e., generate **X**_*I*_ by a given (**X**_*D*_, **X**_*U*_). In GAIN, *G* observes the actual data and generates the needed data according to the observed data. Afterward, *D* takes the completed data from *G*, and tries to discriminate which part of data is generated and which part is actual. Using the collected training data, GAIN will learn the underlying relationships between variables, so that it will gain the capability to accomplish the above-mentioned two tasks. More details of GAIN could be found in Ref. [40].

Based on the trained GAIN model, a personalized lifestyle recommendation approach is then proposed to identify the most suitable lifestyle modification for each risky individual, which consists of two sequential phases: (1) generate applicable alternative lifestyles (Sec. 3.3.2); and (2) select the optimal personalized lifestyle modification. (Sec. 3.3.3**Error! Reference source not found**.).

#### 3.3.2 Phase 1: Generate applicable alternative lifestyles

As demonstrated in Figure 3, this phase consists of two steps, which aims to generate and filter the applicable alternative lifestyles. Specifically, the first step tries to generate various potential alternative lifestyles, i.e.,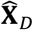, using the trained GAIN model based on the given **X**_*U*_ of each individual. Subsequently, for the second step, the goal is to filter out the inapplicable modifications, since the applicable alternative lifestyles have to satisfy the clinical guidelines (see details in APPENDIX 2). For example, according to Ref. [41, 42], the BMI value for a healthy lifestyle should not be more than 25. Otherwise, the individual should try to reduce the weight. More specifically, increasing the BMI to 25 or higher cannot be treated as an applicable modification. Similarly, if the current BMI is higher than 25, then the suggested BMI cannot be higher than the current value in an applicable modification. Consequently, the generated alternative lifestyles that fit all the clinical guidelines will be selected as applicable lifestyles and the optimal one will be further identified in phase 2.

**Figure 3:**
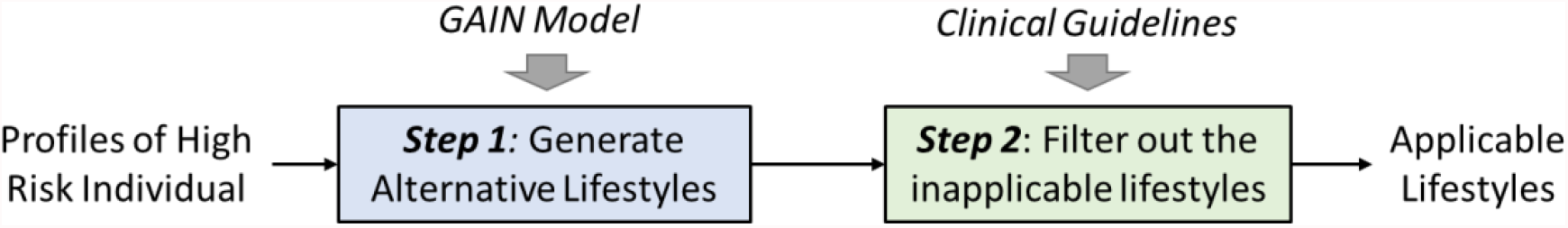
the procedure to generate applicable alternative lifestyles (phase 1).

#### 3.3.3 Phase 2: Identify the optimal personalized lifestyle modification

Phase 2 aims to identify the optimal personalized lifestyle modification, which is the most suitable option for a risky individual by considering the CVD risk reduction, cost from the changes of lifestyle, and uncertainty in disease progression. As shown in Figure 4, this phase consists of three steps: 1) generate replicates (in terms of the indirect changeable variables) for each applicable alternative lifestyles; 2) predict the CVD risk for each replicate; and 3) calculate the expected utility value for each applicable lifestyle and identify the optimal one. Notably, different from the existing studies, the optimal modification in this study is identified by balancing the CVD risk reduction, individual’s preference to change, and disease progression uncertainty, rather than the risk reduction only.

**Figure 4:**
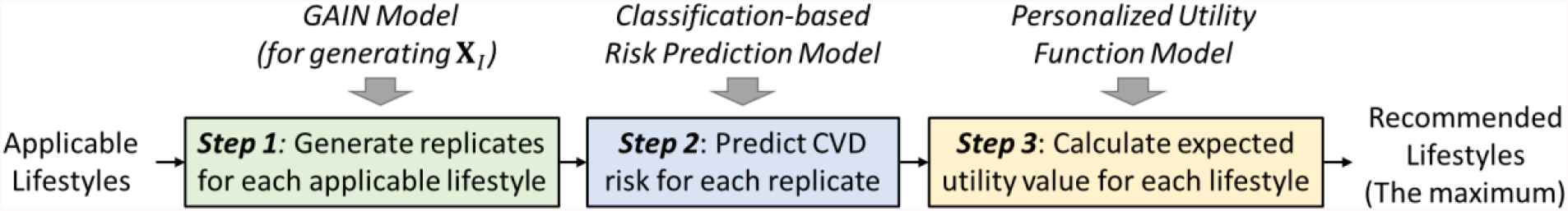
The procedure to identify the optimal personalized lifestyle modification in phase 2.

Step 1 focuses on describing the uncertainty of disease progression under each applicable alternative lifestyle. In practice, the underlying relationship between **X**_*I*_ and (**X**_*D*_, **X**_*U*_) contains significant uncertainties, which needs to be considered when evaluating the suggested alternative lifestyles. Therefore, for each applicable alternative lifestyle generated in phase 1, i.e., given 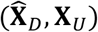, generate *N* replicates for 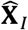 using the GAIN model. Consequently, if *M* applicable alternative lifestyles are generated, each of them can be denoted as 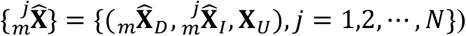, where *m* = 1,2, *⋯, M*.

Subsequently, in step 2, the CVD risk for each 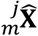 can be further predicted using the trained random forest model 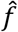 (see more details in Section 3.2), i.e., 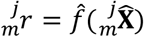. Then for an applicable alternative lifestyle, the predicted overall CVD risk can be described by 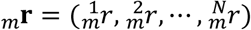, where the uncertainty of disease progression is also quantified in risk estimation.

To further identify the optimal alternative lifestyle for different individuals, besides the overall level of risk, i.e., 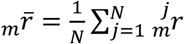, the evaluation should also consider both the uncertainties of CVD risk and the cost of modifications (e.g., quit smoke, reduce BMI, etc.). Thus, it is also needed to develop an effective evaluation metric for decision-making.

To achieve this goal, in step 3, a personalized assessment model, which is based on the expected utility theory, is proposed for optimal alternative lifestyle selection. In the proposed model, assessment is based on a developed personalized regularization-enabled exponential utility function, denoted as *u*(*r*), which is demonstrated in Eq. (4),

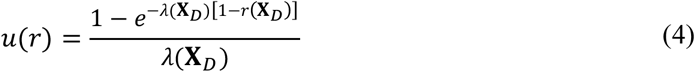

where *λ* is the metric of uncertainty preference (larger *λ* shows less uncertainty averse) and *r* represents the estimated CVD risk for the lifestyle **X**_*D*_. The property and sensitivity of *λ* has been analyzed in [43]. In the traditional exponential utility function, *λ* is a pre-defined constant for each individual. However, in this study, different **X**_*D*_ will lead to different costs of lifestyle modification. Thus, even for the same individual, the value of *λ* may still vary since the cost of lifestyle modification could impact an individual’s preference.

Consequently, the proposed model further extends the constant *λ* to a function of **X**_*D*_, denoted as *λ*(**X**_*D*_), which considers the uncertainties from two aspects, namely, the disease progression (i.e, the variance of *r*) and the cost of lifestyle modifications. The key novel of *λ*(**X**_*D*_) is introducing a personalized regularization term based on the cost of lifestyle modifications. Specifically, given an individual’s original lifestyle 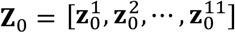, and the cost to change each directly changeable variable, *a*_*i*_, *i* = 1, *…*, 11, (since there are 11 directly changeable variables in **X**_D_, as it is shown in APPENDIX 1) the total cost of lifestyle modification can be written as 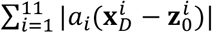. Then by applying this cost to *λ* as a regularization term, *λ*(**X**_*D*_) can be formulated as,

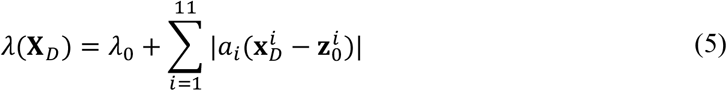

where the first component, *λ*_0_, represents the uncertainty preference of disease progression, which plays the same role as the *λ* in the traditional exponential utility function. The second component, i.e., the proposed regularization term, considers the cost of lifestyle modifications. A large modification cost will increase the value of *λ* and hence reduce the willingness to change.

Then based on the expected utility theory, the optimal 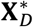 should maximize the expectation of *u*(*r*), i.e., 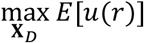. In practice, *E*[*u*(*r*)] can be estimated based on the averaged utility function value of each replicate, i.e., 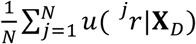. Therefore, the selection of optimal 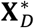 can be formulated by an optimization problem, as shown in Eq. (6),

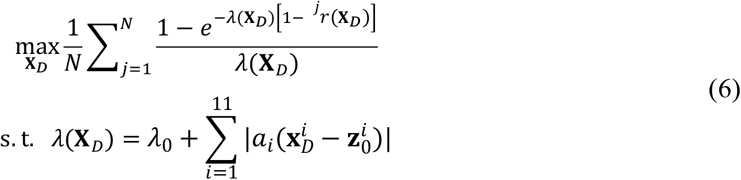

where ^*j*^*r*(**X**_*D*_) represents the predicted disease risk for the *j*th replicate under lifestyle **X**_*D*_. For a specific individual, from the *M* applicable alternative lifestyles (each has *N* replicates) generated from the GAIN model, the optimal 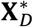 can be thereby selected through Eq. (6).

### 3.4 Performance validation

For the proposed method, it is also critical to validate its effectiveness. However, it is challenging since there is no actual patients are tested with a comparison target. Therefore, alternatively, this study aims to conduct the performance validation through three aspects: (1) the classification accuracy for risk prediction (Sec. 3.4.1**Error! Reference source not found.Error! Reference source not found**.); (2) the effectiveness of the generated indirectly changeable variables (Sec. **Error! Reference source not found.Error! Reference source not found**.); and (3) the effectiveness of the recommended alternative lifestyles for risk reduction (Sec. **Error! Reference source not found.Error! Reference source not found**.).

#### 3.4.1 Validation for the classification-based risk prediction

To validate the classification performance, the data are randomly separated into two parts, one training set (50%) and one testing set (50%). The classifiers, including the RF, SVM, and k-NN, are first trained using the training set, and then the testing set is used for validation. The classification performances are measured by the receiver operating characteristic (ROC) curve with its AUC score.

#### 3.4.2 Validation for the effectiveness of the generated indirectly changeable variables

Validating the effectiveness of generated indirectly changeable variables also plays a very important role in this study. As demonstrated in Figure 5**Error! Reference source not found**., the proposed validation framework will first select samples from the testing set and then use the trained GAIN model to generate the indirectly changeable variables 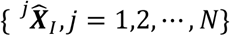 for each sample with *N* replicates conditional on the actual lifestyle **X**_*D*_ and characteristics **X**_*U*_. Subsequently, the corresponding risk { ^*j*^*r, j* = 1,2, *⋯, N*} can be thereby predicted. If { ^*j*^*r*} is consistent with the risk that is predicted using the real observations of indirectly changeable variables, i.e., *r*_*a*_, through comparison, then it can be concluded that the generated 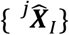 is effective. Specifically, the consistency comparison between { ^*j*^*r*} and *r*_*a*_ can be conducted through two aspects: (1) to compare if *r*_*a*_ is inside the range of { ^*j*^*r*}; and (2) for all the samples, to compare if the average of { ^*j*^*r*} is highly correlated with the corresponding *r*_*a*_.

**Figure 5:**
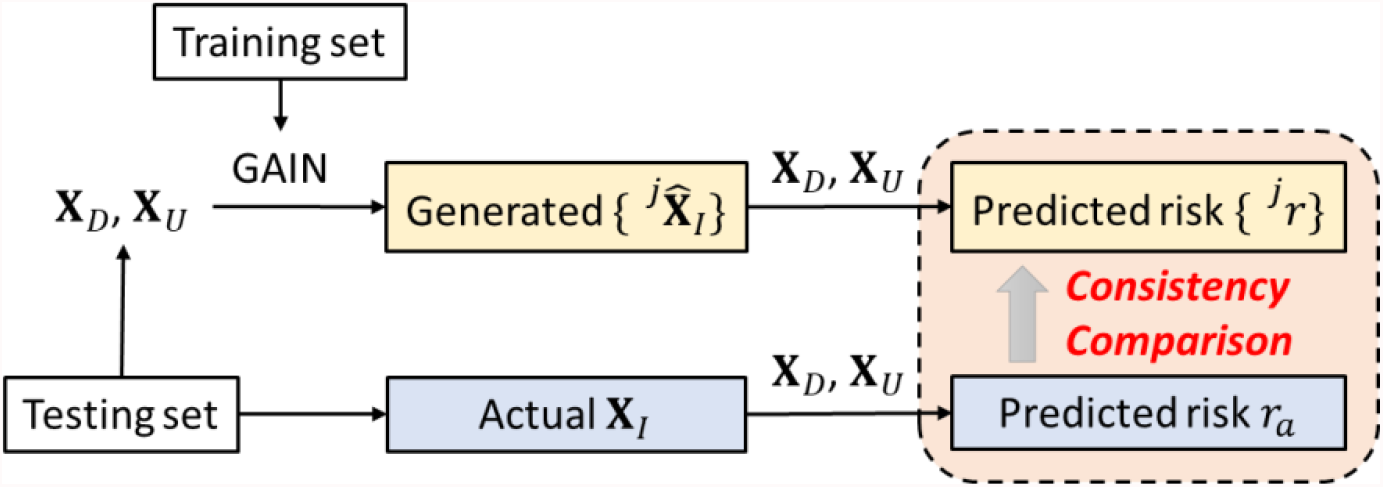
The proposed framework to validate the effectiveness of the generated indirectly changeable variables.

#### 3.4.3 Validation for the effectiveness of the recommended lifestyle modification

Another critical validation in this study is the effectiveness of the recommended lifestyle modification for each individual. Essentially, the ultimate goal is to validate if the recommended lifestyle can significantly reduce CVD risk. Mathematically, it can be formulated as a hypothesis testing problem, i.e.,

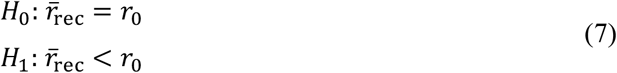

where *r*_0_ represents the original CVD risk and 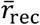 represents the mean CVD risk after the recommended lifestyle modification. For a specific individual, *r*_0_ can be predicted from his/her original lifestyle factors using the classification models developed in Sec. 3.2. The mean CVD risk after the recommended lifestyle modification can be inferred by generating multiple replicates of patient’s indirectly changeable variables from the GAIN model and quantifying their risks, which are denoted as { ^*j*^*r*_rec_}. Then the hypothesis in Eq. (7) can be tested through *t*-test.

As demonstrated in Figure 6, the validation can be summarized by three steps: (1) pick up the high-risk individuals from the testing set; (2) predict the risk based on their current lifestyle and recommended lifestyle modification (with replicates), respectively; and (3) perform *t* test to see if 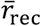 is significantly lower than *r*_0_.

**Figure 6:**
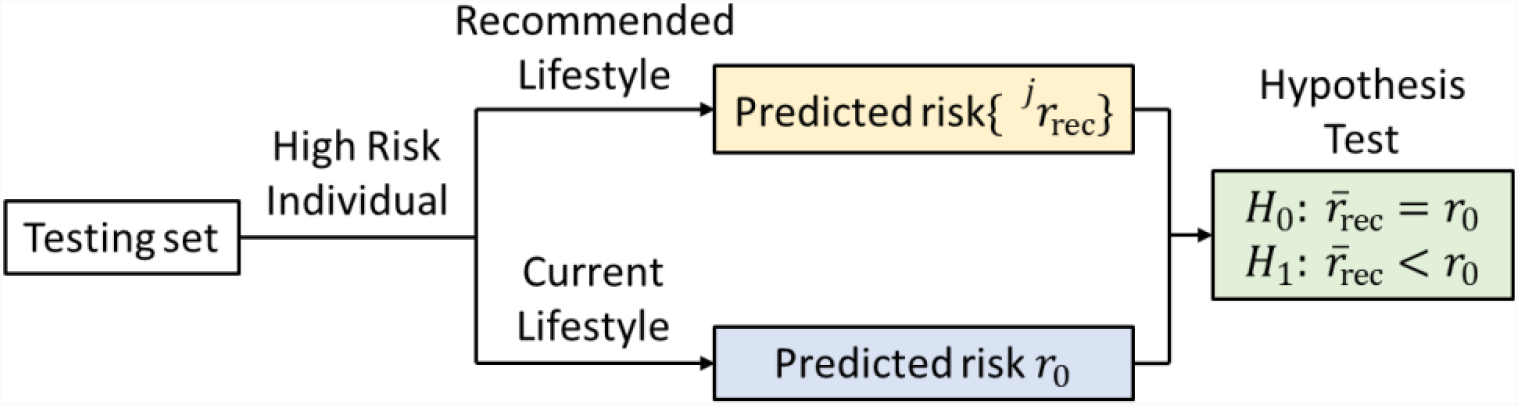
The proposed framework to validate the effectiveness of the recommended lifestyle.

## 4 Results

Based on the methodology proposed in Sec. 3, this section presents the validation results based on the actual data selected from the ARIC database. The classification results are presented in Sec. **Error! Reference source not found**.. Sec. **Error! Reference source not found**. demonstrates the effectiveness validation for the generated indirectly changeable variables and followed by the validation results of the recommended lifestyle medication for the high-risk individuals (Sec. 4.3**Error! Reference source not found**.).

### 4.1 Classification results

As presented in Sec. 3.4.1, this study randomly selected 50% of samples in the dataset for training, and the other 50% are used for testing. Then based on the ROC curve and AUC score, the comparison between three popular classification methods, i.e., random forest, SVM, and *k*-NN classifiers was conducted. As presented in Figure 7, it can be observed that random forest can provide superior classification performance than the other two methods for all outcome variables. Therefore, the random forest classifier is selected in this study for CVD risk prediction.

**Figure 7:**
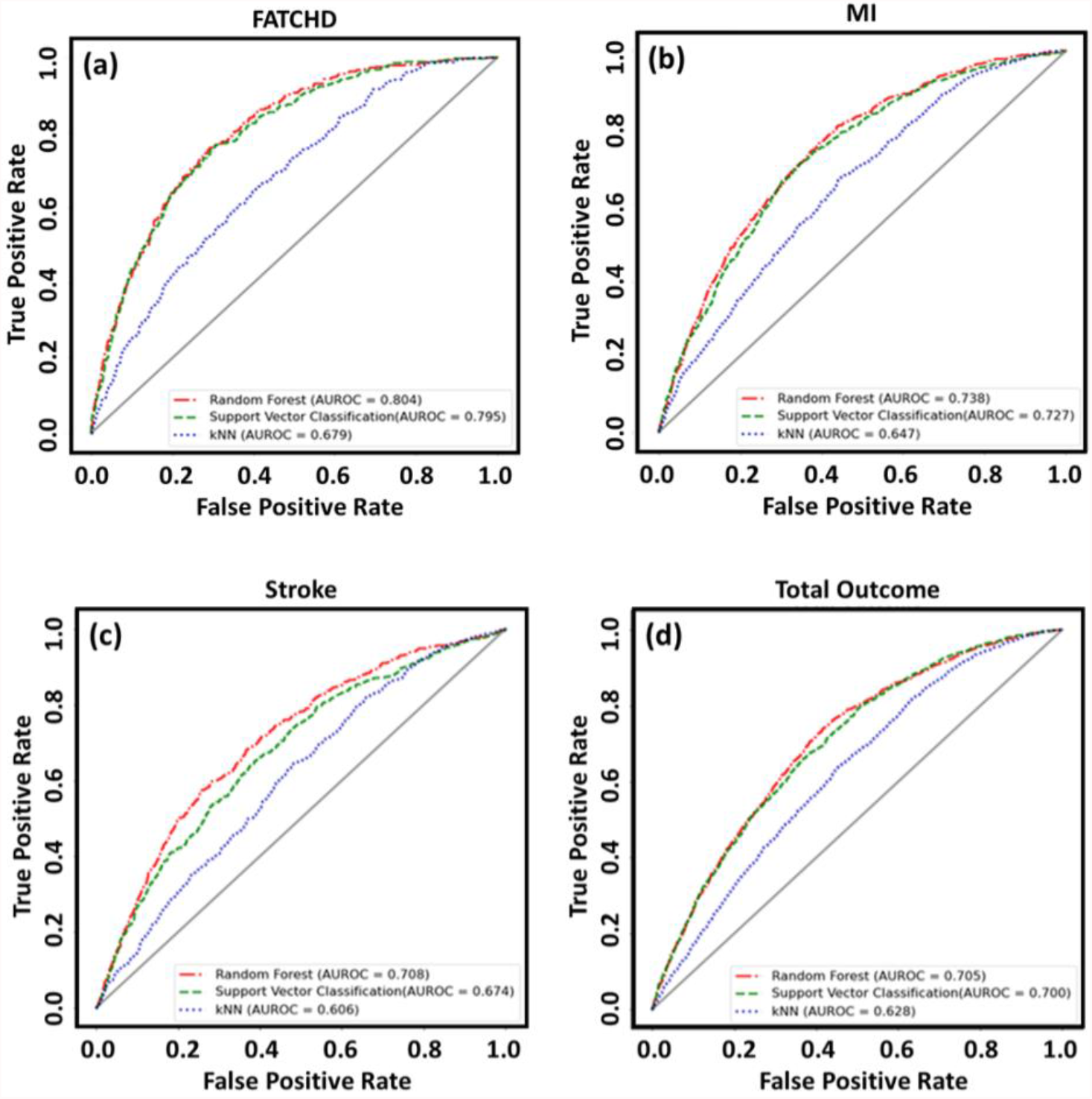
ROC Curves based on different classification models for the four outcome variables. (a) Fatal CHD; (b) MI; (c) Stroke; and (d) Total outcome.

Furthermore, the detailed classification results using random forest are presented in **Error! Reference source not found**.. For all outcome variables, the AUC score can achieve at least 70%, which indicates that the trained random forest model 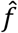 is able to effectively predict the CVD risk of an individual based on its profile (**X**_*U*_, **X**_*I*_, **X**_*D*_). Besides, 10-folder cross validation is also applied in this study, and the AUC scores also demonstrate similar results.

**Table 1:** Data distributions and AUC Scores of the Outcomes

| CVD Type | Sample Size | Diagnosed Patient Size | Diagnosed Patient Rate | AUC Score | AUC Score (Cross Validation) |
| --- | --- | --- | --- | --- | --- |
| Fatal CHD | 10744 | 677 | 0.063 | 0.82 | 0.81 |
| MI | 11635 | 1538 | 0.132 | 0.74 | 0.72 |
| Stroke | 11292 | 1195 | 0.106 | 0.70 | 0.70 |
| General Outcome | 13654 | 3557 | 0.260 | 0.70 | 0.69 |

### 4.2 Evaluation of the generated indirect changeable variables

This section presents the evaluation results for the generated indirectly changeable variables. To conduct the validation, 200 of the truly healthy individuals who are correctly classified as healthy, and 200 of the truly unhealthy individuals who are correctly classified as unhealthy are selected from the testing set (50% samples, see Sec. 4.1). For the training of GAIN, the batch size and the number of iterations are set as 32 and 5000, respectively. As presented in Sec. 3.4.2, for each individual, the indirectly changeable variables with *N* replicates, i.e., 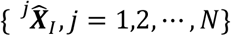, were generated conditional on the individual’s actual lifestyle ***X***_*D*_ and characteristics ***X***_*U*_. Then the corresponding risk { ^*j*^*r, j* = 1,2, *⋯, N*} for each replicate was predicted as well. Notably, the number of replicates (*N*) is set to 500. As presented in Figure 8a and Figure 8c, in terms of the Fatal CHD and Stroke, the Pearson correlation coefficients between the averaged { ^*j*^*r*} and the corresponding *r*_*a*_ can achieve 91% and 95%, respectively. Furthermore, based on MI and the total outcome, the correlation cofficients are also signigicantly higher than 80% (see Figure 8b and Figure 8d). Thus, the results indicate that the synthesized indirectly changeable variables using GAIN can well approximate to actual situation.

**Figure 8:**
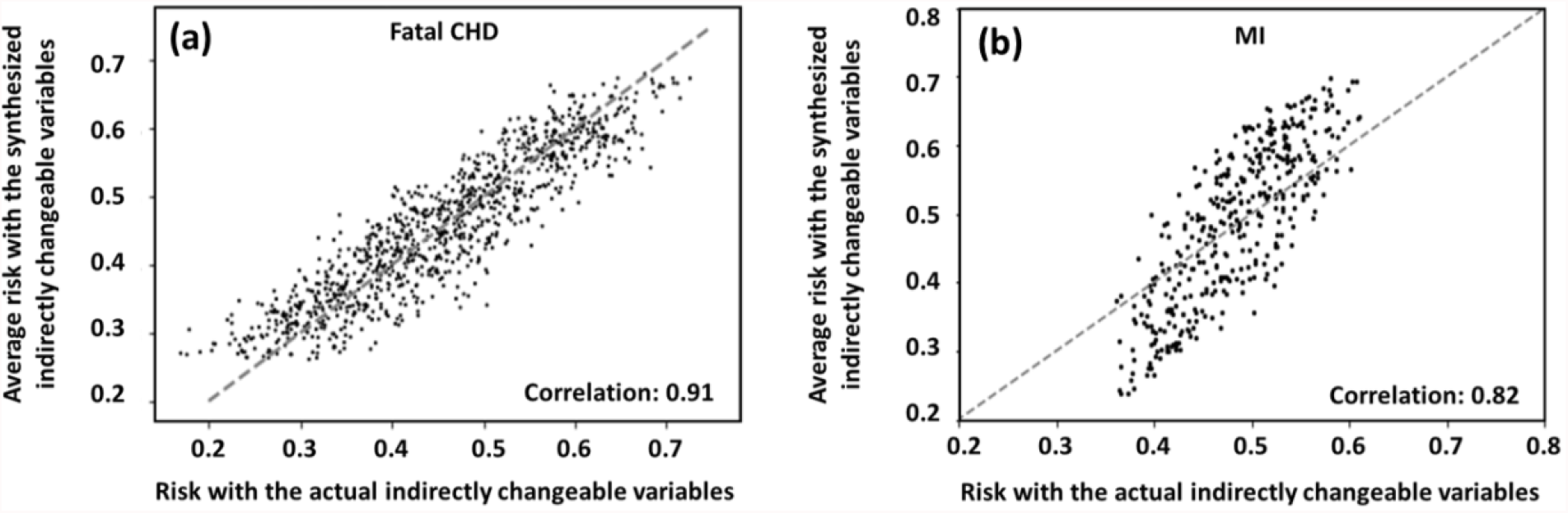
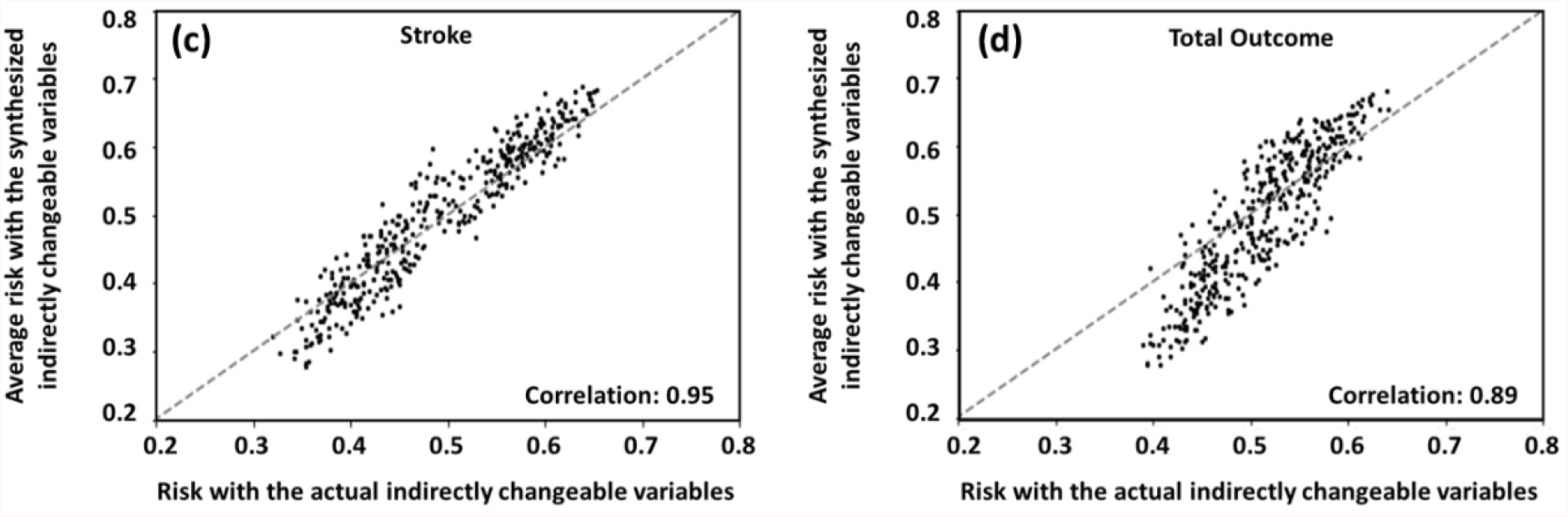
The correlation between average risk with the synthesized indirectly changeable variables vs. risk with actual indirectly changeable variables. (a) Fatal CHD; (b) MI; (c) Stroke; (d) Total outcome.

Moreover, 10 individuals were randomly selected from the data to be shown in Figure 9, and then further compare if *r*_*a*_ is inside the range of { ^*j*^*r*} for each individual, in terms of the Fatal CHD. As demonstrated in Figure 9, the light blue boxes represent the range of { ^*j*^*r*}, and the dark blue line inside each box represents the value of *r*_*a*_. From the results it can be observed that the range of the { ^*j*^*r*} can well cover the value of *r*_*a*_, which further indicates that the GAIN-based generation for **X**_*I*_ is effective.

**Figure 9:**
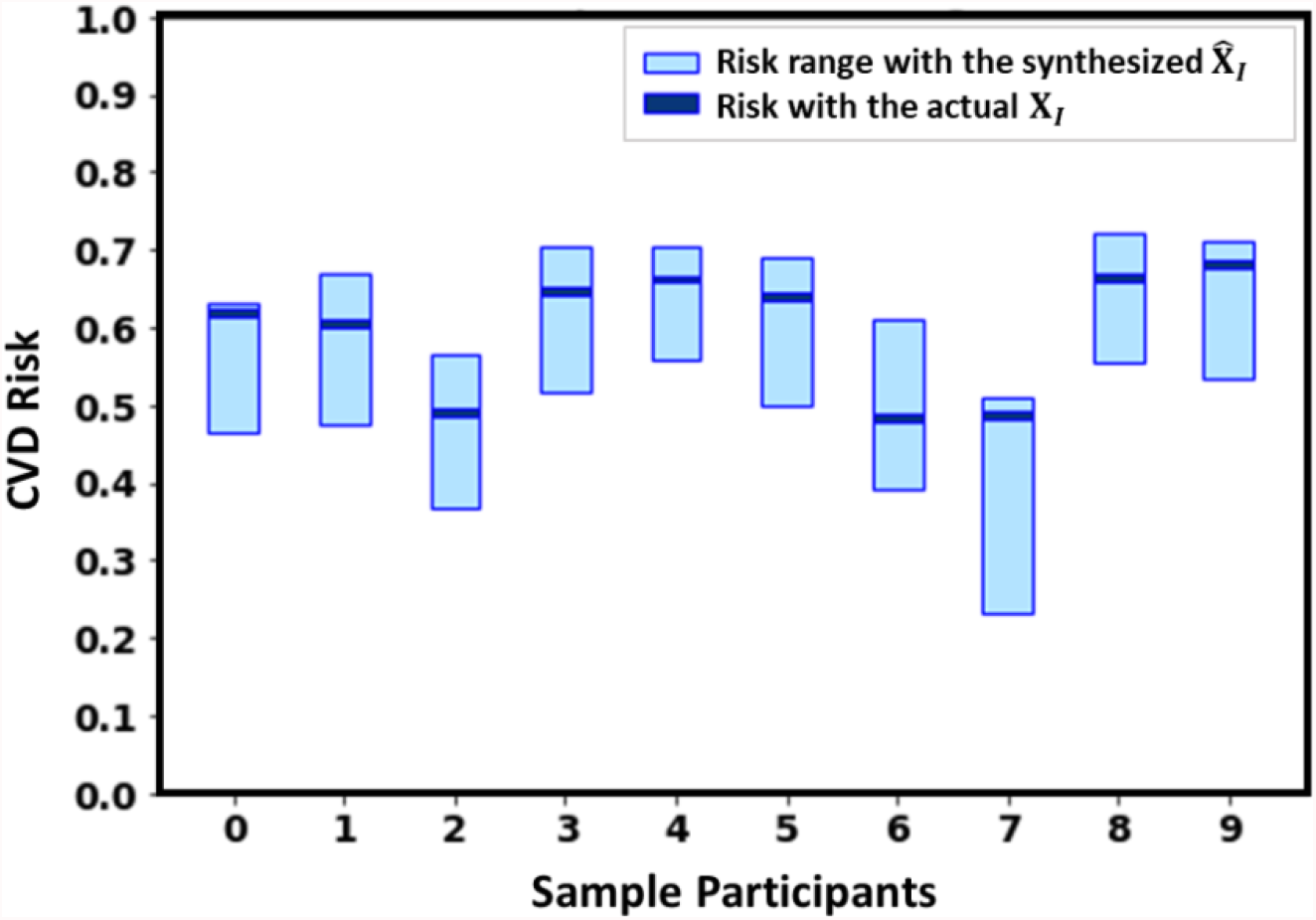
Risk ranges of the replicates after synthesizing the indirectly changeable features by GAIN and the original risk with original indirectly changeable features for 10 randomly selected participant.

### 4.3 Evaluation of the suggested lifestyle modifications

As introduced in Sec. 3.4.3, this section demonstrates the validation results for the recommended lifestyle modifications using hypothesis test. To avoid the potential bias in invalidation, we only choose the truly unhealthy individuals who are correctly classified as unhealthy from the testing set (50% samples, see Sec. 4.1) which indicates 285, 644, 513, 1492 in terms of fatal CHD (*y*_1_), MI (*y*_2_), stroke (*y*_3_), and total outcome (*y*_4_), respectively. To identify the optimal lifestyle modification for all of these individuals, the number of replicates (*N*) for each applicable modification is set to 500. Regarding the parameters in the proposed utility function model, without loss of generality, we set *λ*_0_ = 0 (indicate low risk aversion) and **a** = (*a*_1_, *a*_2_, *⋯, a*_11_) = (1, 0.05, 0.1, 0.1, 0.05, 0.1, 0.1, 0.05, 0.05, 0.05, 0.05) × 10^2^. In practice, based on different individual cases, these personalized parameters could be adjusted flexibly.

For fatal CHD (*y*_1_), the hypothesis test results in terms of *p*-value are presented in Figure 10, which shows that the *p*-values for 98% individuals are lower than 0.05, and only 1.6% are larger than 0.1. The results for MI (*y*_2_), stroke (*y*_3_), and total outcome (*y*_4_) are presented in Figure 11, Figure 12, and Figure 13, respectively, which also show similar patterns. Therefore, the validation results demonstrate the proposed method can significantly reduce the potential CVD risk of risky individuals.

**Figure 10:**
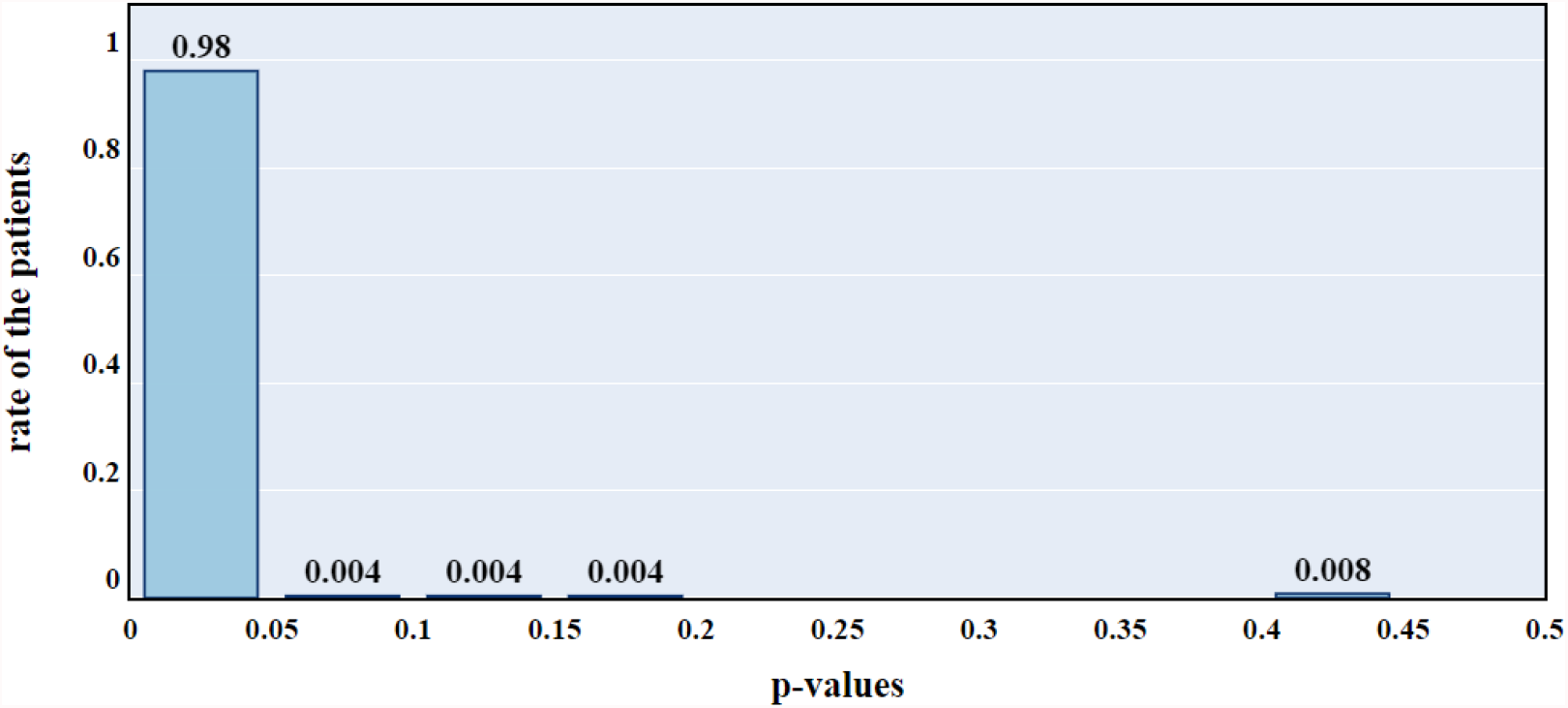
Hypothesis testing results for fatal CHD risk reduction based on recommended lifestyle modification.

**Figure 11:**
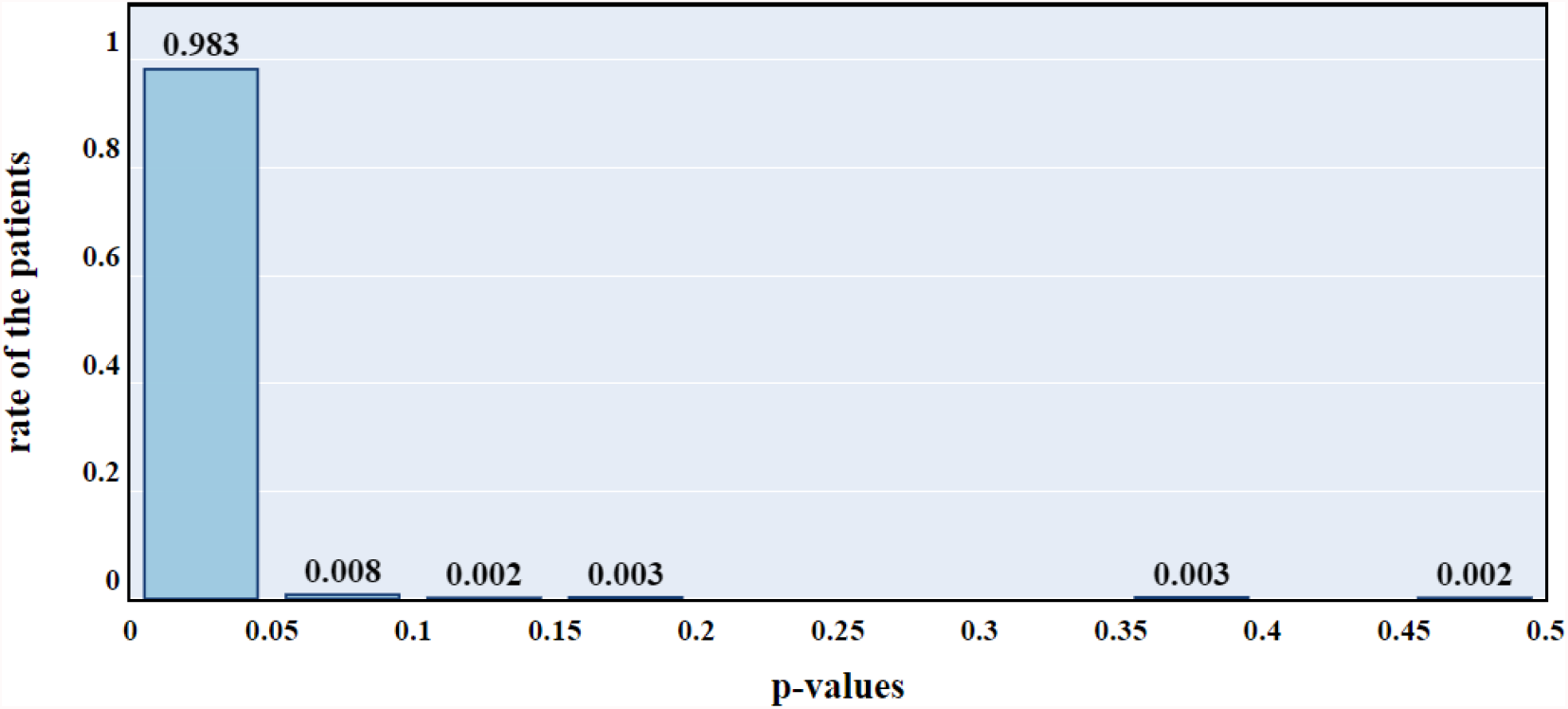
Hypothesis testing results for MI risk reduction based on recommended lifestyle modification.

**Figure 12:**
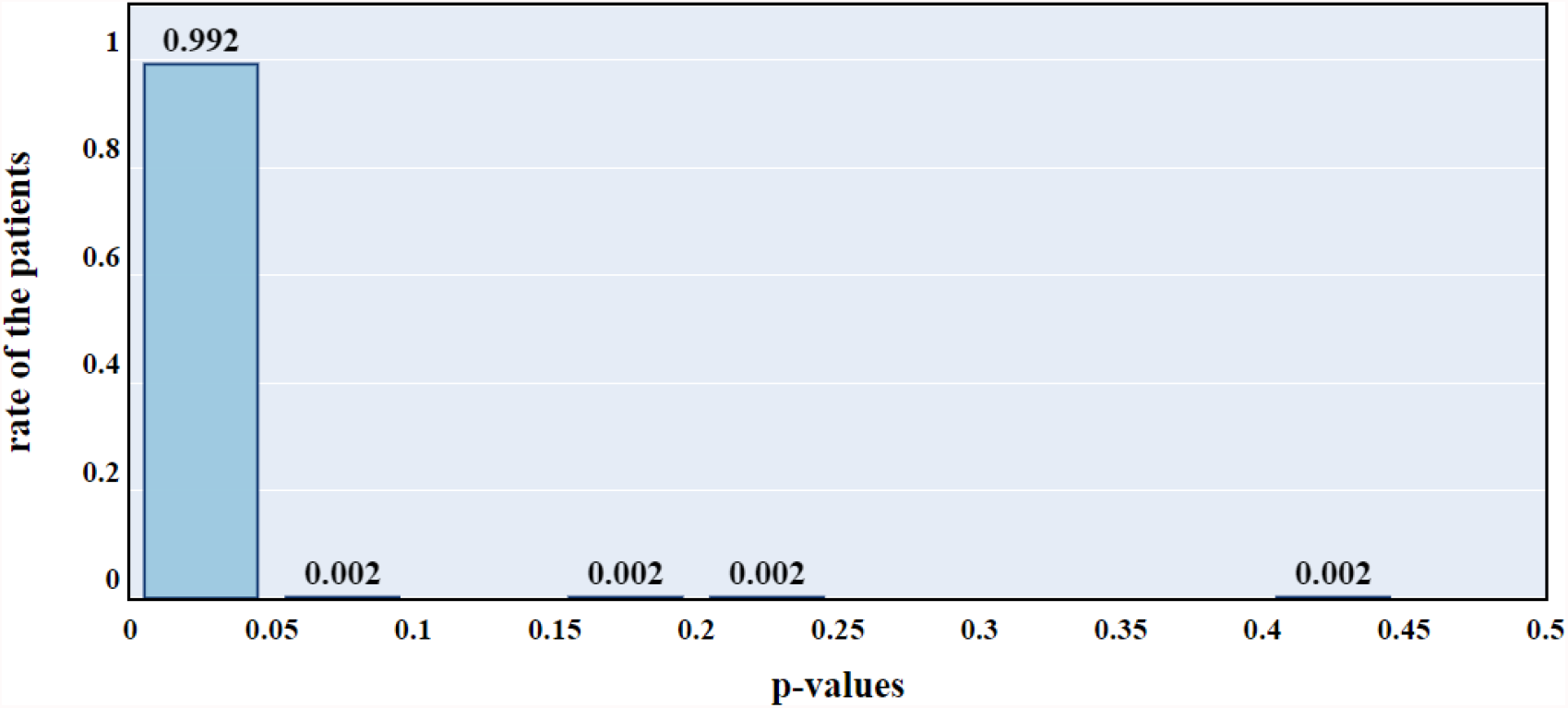
Hypothesis testing results for stroke risk reduction based on recommended lifestyle modification.

**Figure 13:**
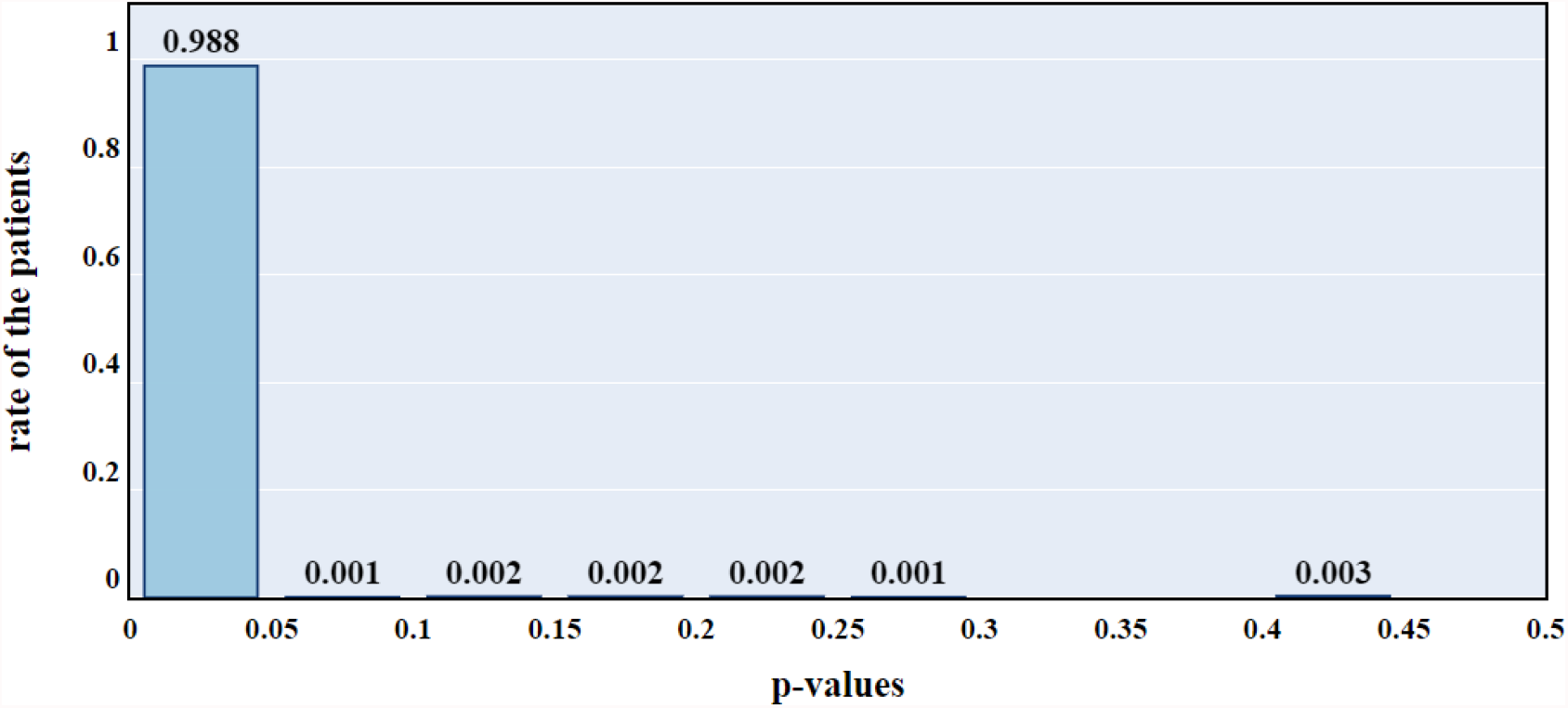
Hypothesis testing results for overall CVD risk (i.e., the total outcome) reduction based on recommended lifestyle modification.

Specifically, the average risk reduction of fatal CHD can achieve 9.4% (with the standard deviation 5.19%), after applying the recommended lifestyle modfication. Similarly, the average risk of diagnosing with MI and stroke in 10 years can be also reduced by 8.65% (with the standard deviation 4.58%) and 5.3% (with the standard deviation 2.93%), respectively. Based on the overall CVD outcome, the proposed method is able to provide 5.7% reduction (with the standard deviation 3.29%) in the risk of diagnosing with a CVD type (fatal CHD, MI, or Stroke) in 10 years.

Figure 14 illustrates the recommended lifestyle modification for a randomly selected high risk individual to reduce the risk of fatal CHD. It suggests a reduction in saturated fatty acid (g/day) and cholesterol (mg), while keeping the BMI, alcohol (g/day), dietary fiber, total fat (g/day), and smoking status the same. Besides, it also recommends to increase the physical activity (h/week), protein intake (g), total energy (mg/dL), and carbohydrate (g). In other words, this suggested modification provided by the proposed method mainly recommends this individual to adjust the source of energy, i.e., to replace the energy taken from the saturated fatty acid and cholesterol by the energy from other carbohydrates, protein. With the suggested lifestyle modification, the risk of diagnosing with fatal CHD in 10 years can be reduced by 19%.

**Figure 14:**
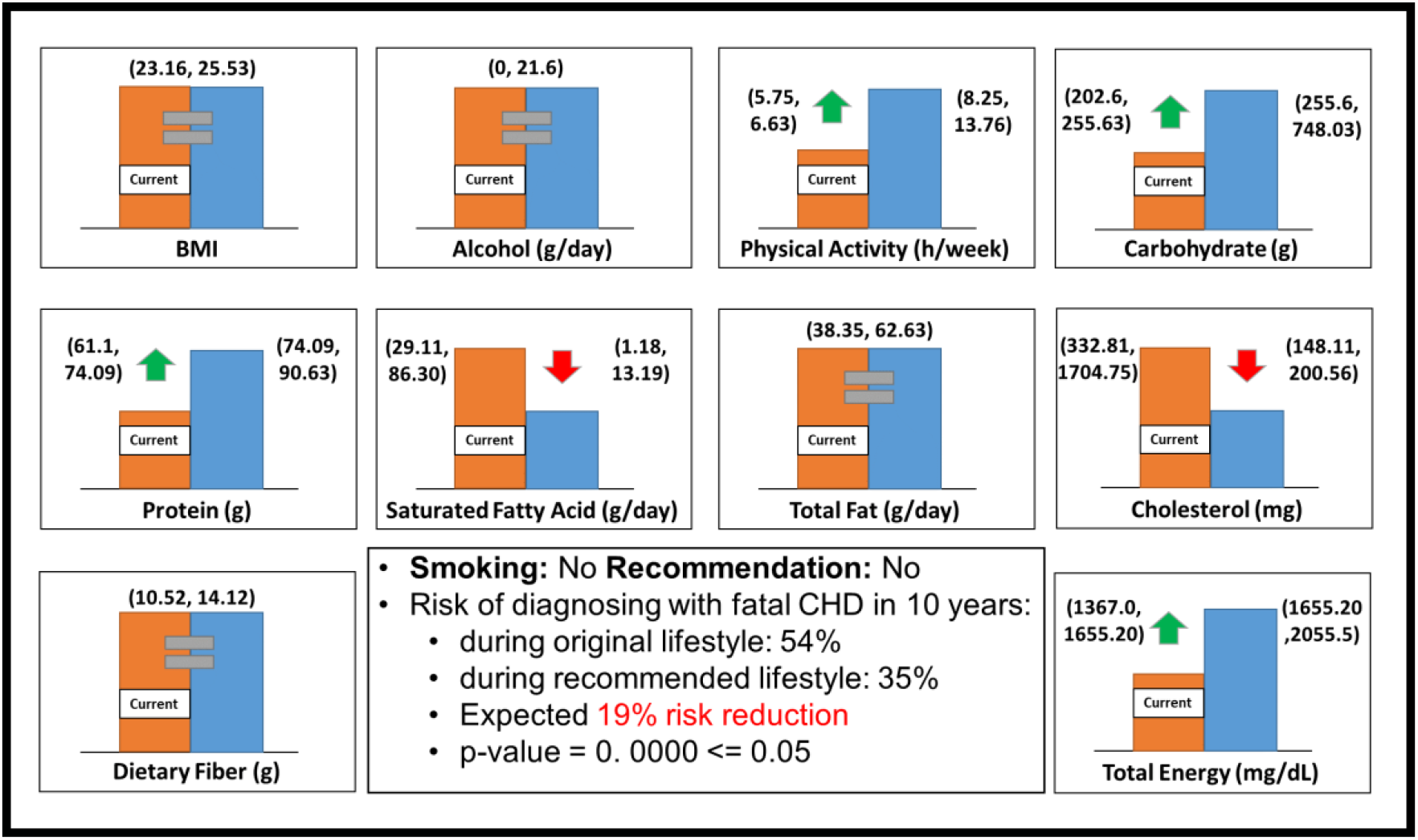
Lifestyle recommendations and its performance for one of the randomly selected participants.

## 5 Conclusions and Discussions

This research established a data-driven personalized lifestyle recommendation algorithm for CVD risk reduction by incorporating the random forest classification, GAN, and utility function model. The proposed method has significant advantages over the existing personalized lifestyle recommendation algorithms by explicitly capturing the complex interactions between risk factors, uncertainty of disease progression under lifestyle modifications, as well as patients’ willingness and cost to change lifestyle behaviors. In the proposed method, the GAN-based model learns the patterns of healthy individuals’ lifestyles and the complex interactions between variables. Therefore, the applicable healthy lifestyle modifications that are consistent with clinical guidelines can be identified, and their associated changes in indirectly changeable variables can be predicted as well, which can indicate the diease progression more accurately. The regularized utility function model proposed in this study provides a quantitative approach to evaluate the utility of lifestyle modifications in practice by balancing the effectiveness in risk reduction, uncertainty in disease progression, and the patient’s preferences. Therefore, the personalized lifestyle modification recommended from the proposed algorithm has great potential to effectively reduce the risk of common CVDs for each patient with sufficient considerations of patient’s likelihood to adhere the recommendation.

To conduct this proof-of-concept study, the patient-level data from the ARIC database is utilized for model training and validation, since the dataset includes a fairly large number of participants and extensive measured variables that are related to occurance of common CVDs. Through the proposed method, patients’ CVD risk could be effectively assessed and the invidualized optimal lifestyle modification could be timely updated. Meanwhile, based on the proposed novel utility function model, the recommendation provided by the proposed method can also well balance the potential of risk reduction and the individual’s preferences. For example, some high risk patients are recommended to reduce the saturated fatty acid and cholesterol in dietary and increase the physical activity and protein intake for developing healthier lifestyle behaviors (see Figure 14). The recomemdations are consistent with the evidences found in existing literature [1, 5, 6, 12]. Thus, our methodology could make great contributions to the clinical practice for CVD prevention. In practice, the proposed methodology can be applied as a decision-support tool for healthcare professionals and high-risk individuals in conducting an optimal applicable lifestyle by learning from the related healthcare data.

In the potential real-world applications, to make the recommendation scenario more realistic, we can further consider more CVD-related risk factors. For example, the socio-economic factors, including fast food options, availability of fresh and healthy food, time availability to increase exercise timing, etc. With more related variables, it has potential to further increase the accuracy of our proposed method. In addition, the genetic factors such as the family genetic history might be also helpful to improve the recommendation performance. It is also worth mentioning that jointly considering the data from multiple visits may also be helpful. For example, the year-by-year longitudinal patterns, such as the change of diet habits and medication history, could be learned by the machine learning algorithms and utilized for offering better recommendation options. Therefore, the machine learning algorithms that are capable of handling the longitudual patterns could be adopted in the future work.

In summary, this study developed a data-driven personalized lifestyle recommendation framework for CVD prevention. A proof-of-concept case study based on the ARIC dataset has demonstrated that our method is able to provide appropriate individualized lifestyle recommendation and thereby effectively reduce the CVD risk. It shows the potential to enable better use of individual-level healthcare data for more effective CVD prevention. Furthermore, this study is also very promising to be extended to more types of diseases (e.g., hypertension, diabetes, etc.) and can make more contributions to the researches related to behavior change, personalized treatment, and medicine. In the future work, more related datasets and emerging machine learning techniques will be incorporated to further improve the recommendation accuracy and capability.

## Data Availability

All data produced in the present study are available upon reasonable request to the authors.

## APPENDIX 1

**Table 2:**
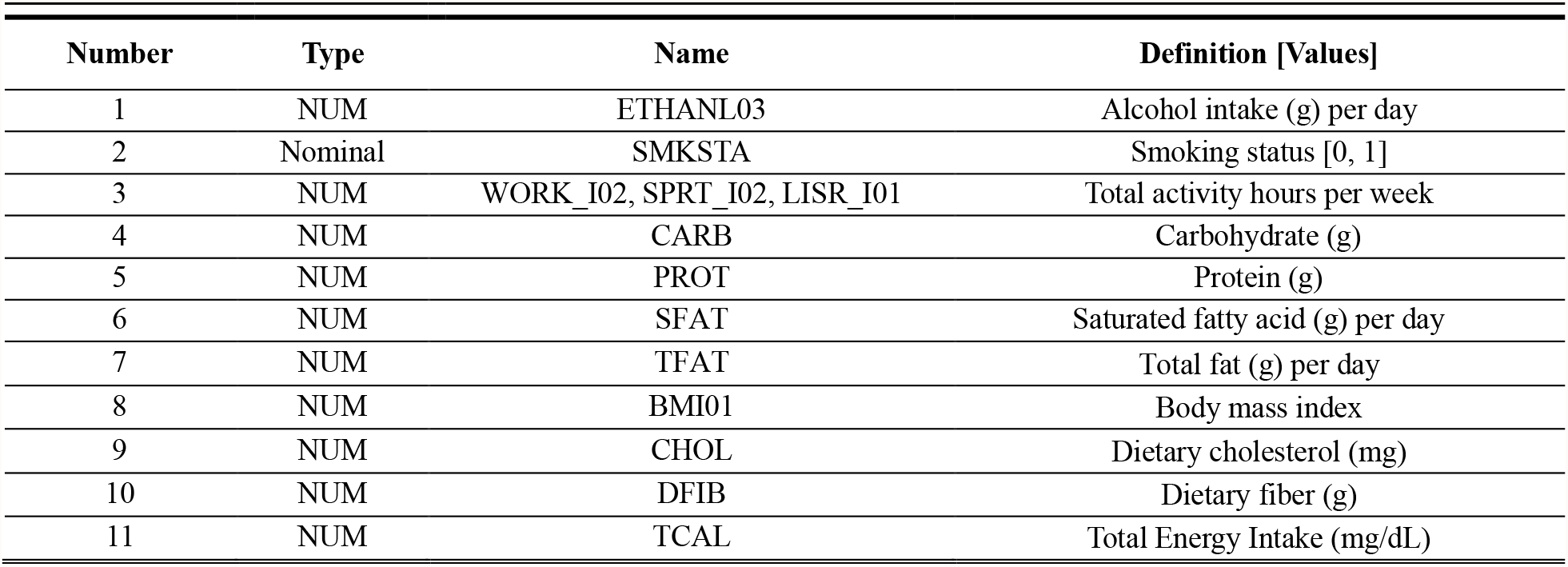
The list of the directly changeable features

| Number | Type | Name | Definition [Values] |
| --- | --- | --- | --- |
| 1 | NUM | ETHANL03 | Alcohol intake (g) per day |
| 2 | Nominal | SMKSTA | Smoking status [0, 1] |
| 3 | NUM | WORK_I02, SPRT_I02, LISR_I01 | Total activity hours per week |
| 4 | NUM | CARB | Carbohydrate (g) |
| 5 | NUM | PROT | Protein (g) |
| 6 | NUM | SFAT | Saturated fatty acid (g) per day |
| 7 | NUM | TFAT | Total fat (g) per day |
| 8 | NUM | BMI01 | Body mass index |
| 9 | NUM | CHOL | Dietary cholesterol (mg) |
| 10 | NUM | DFIB | Dietary fiber (g) |
| 11 | NUM | TCAL | Total Energy Intake (mg/dL) |

**Table 3:** The list of indirectly changeable features

| Number | Type | Variable | Definition [Values] |
| --- | --- | --- | --- |
| 1 | NUM | HDLSIU02 | HDL cholesterol in mg/dl |
| 2 | NUM | LDLSIU02 | LDL cholesterol in mg/dl |
| 3 | NUM | TCHSIU01 | Total cholesterol in mmol/L |
| 4 | NUM | TRGSIU01 | Total triglycerides in mmol/L |
| 5 | NUM | SBPA21 | 2nd and 3rd Systolic blood pressure average |
| 6 | NUM | SBPA22 | 2nd and 3rd Diastolic blood pressure, blood pressure average |
| 7 | NUM | ANTA07A | Waist girth to nearest CM |
| 8 | NUM | ANTA07B | Hip girth to nearest CM |
| 9 | NUM | ECGMA31 | Heart rate |
| 10 | NUM | HMTA03 | White blood count |
| 11 | NUM | APASIU01 | Apolipoprotein AI (MG-DL) |
| 12 | NUM | APBSIU01 | Apolipoprotein B (MG-DL) |
| 13 | NUM | LIPA08 | APOLP (A) DATA (UG-ML) |
| 14 | NUM | CHMA09 | Creatinine (MG-DL) |

**Table 4:** The list of unchangeable features

| Number | Type | Variable | Definition [Values] |
| --- | --- | --- | --- |
| 1 | NUM | CIGTYR01 | Cigarette years of smoking |
| 2 | Nominal | ELEVEL01 | Education level [(1) grade school or 0 years education, (2) high school, but no degree, (3) high school graduate (4), vocational school, (5) college (6) graduate school or professional school] |
| 3 | Nominal | GENDER | Sex |
| 4 | Nominal | RACEGRP | Race |
| 5 | NUM | V1AGE01 | Age |
| 6 | Nominal | DIABTS02 | Diabetes [0, 1] |
| 7 | Nominal | HYPERT04 | Hypertension [0, 1] |
| 8 | Nominal | HYPTMDCODE01 | Hypertension medication in the past 2 weeks |
| 9 | Nominal | CHOLMDCODE01 | Cholesterol-lowering medication use [0, 1] |
| 10 | NUM | ANTA01 | Standing height to nearest CM |
| 11 | Nominal | CIGT01 | Smoking status history |
| 12 | NUM | HYPTMD01 | HYPERTENSION LOWERING MED. USE |
| 13 | NUM | ANTICOAGCODE01 | Used Anticoagulates (at Visit 1) last 2 weeks (0=No, 1=Yes) based on 2004 Med Code |
| 14 | NUM | ASPIRINCODE01 | Used Aspirin-containing analgesics (at Visit 1) in last 2 weeks (0=No, 1=Yes), based on 2004 Med Code |
| 15 | NUM | STATINCODE01 | Used Statin (at Visit 1) last 2weeks (0=No, 1=Yes) based on 2004 Med Code |

## APPENDIX 2

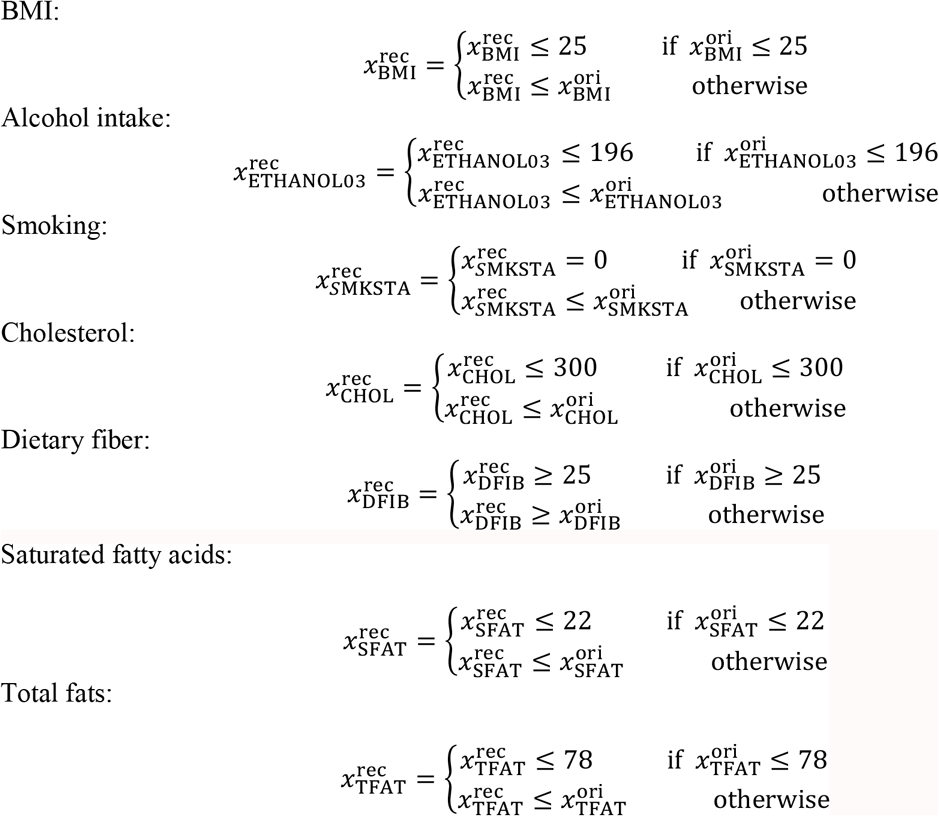

